# High-risk Molecular Features Eclipse Genomic Complexity in Predicting CLL Patient Outcomes; Insights from the UK CLL4, ARCTIC and ADMIRE Trials

**DOI:** 10.1101/2025.07.22.25331923

**Authors:** H Parker, L Carr, K Norris, A Nilsson-Takeuchi, B Stevens, H Amarasinghe, Latha Kadalayil, M Else, A Pettitt, P Hillmen, A Schuh, R Walewska, DM Baird, DG Oscier, C Pepper, D Bryant, J Gibson, JC Strefford

**Author notes:** Correspondence: Prof. Jonathan C Strefford, Faculty of Medicine, University of Southampton, Cancer Genomics Group, MP824 Somers Building, Southampton General Hospital, Tremona Road, Southampton, SO16 6YD, UK. First authorship (H Parker, L Carr). last authorship (D Bryant, J Gibson, JC Strefford). The authors declare no potential conflicts of interest.

## Abstract

High genomic complexity is linked to poor prognosis in chronic lymphocytic leukaemia (CLL), but its independent prognostic value remains uncertain amid emerging biomarkers.

We analysed copy number alterations (CNA) in 495 treatment-naïve patients from three randomized trials (CLL4, ADMIRE, ARCTIC), incorporating IGHV status, telomere length (TL), targeted sequencing, and DNA-methylation subtypes. Patients harboured low (LGC, 0–2 CNAs; n=334), intermediate (IGC, 3–4 CNAs; n=97), or high (HGC, ≥5 CNAs; n=64) genomic complexity.

U-CLL (81%, p<0.001) and short TL (61%, p<0.05) were enriched in HGC, and TL inversely correlated with CNA burden (τ = –0.147, p<0.001). 62% of HGC patients were n-CLL. TP53 dysfunction was associated with HGC (36%, p<0.001). Trisomy 12 and NOTCH1 mutations, were enriched in LGC (p<0.001). HGC predicted shorter progression-free and overall survival in all univariate models but only remained independently prognostic for OS only in CLL4 (HR=1.61, p=0.02). Of 64 HGC patients, 23 had TP53 dysfunction; 92% of TP53 wild-type cases had other high-risk features (TL-S, U-CLL, or n-CLL).

HGC is associated with adverse outcomes but may reflect underlying biological risk rather than serve as an independent biomarker. Its interplay with telomere attrition, immunogenetics, and epigenetic subtype warrants further validation in targeted therapy-treated cohorts.

## Introduction

Chronic lymphocytic leukaemia (CLL) is a heterogeneous disease characterized by significant biological and clinical variability, affecting disease progression, treatment response, and patient outcomes. Key prognostic indicators, such as TP53 aberrations and unmutated IGHV genes (U-CLL), are essential for risk stratification but do not include all patients at risk of poor survival. As CLL cells proliferate in the bone marrow and lymph nodes, some clones develop complex genomes marked by structural lesions, aneuploidy, and somatic mutations. Genomic complexity (GC) is defined by chromosomal alterations: high-GC (HGC) is defined by five or more karyotypic or copy number alterations (CNAs), while GC encompasses three or more. Notably, although TP53 aberrations are often associated with genomic instability, over 20% of GC cases lack detectable TP53 lesions, indicating that other factors may contribute to GC, and certain chromosomal changes like trisomy 12 and trisomy 19 may positively or neutrally impact clinical outcomes (1).

Recent studies have linked HGC to poorer prognosis (2–5) and an increased risk of Richter’s transformation (6), often associating with negative indicators like TP53 disruption and unmutated IGHV CLL (U-CLL) (7, 8). In the context of traditional chemotherapy, chlorambucil-based regimens showed improved overall survival (OS) for patients without GC compared to those with HGC (9). The CLL14 trial indicated that HGC patients had lower response rates and significantly shorter progression-free survival (PFS), and OS compared to patients without GC (10). For targeted treatments, while some data suggest shorter OS for HGC patients treated with ibrutinib (11), other studies reported high overall response rates, though with shorter durations and poorer PFS (12). The MURANO study revealed that HGC patients on venetoclax had lower rates of undetectable minimal residual disease (uMRD) and shorter PFS (13), which the CLL13 trial confirmed (14). Interestingly, the CLL14 trial found no significant differences in response rates or survival between HGC and non-GC patients (10). Similarly, treatment with the PI3K-delta inhibitor, idelalisib, showed no significant difference in response rates for HGC patients (15).

Additionally, HGC often coexists with telomere length (TL) attrition (16). Critical TL shortening leads to uncapped telomeres, driving fusion events and accumulating genetic lesions like del(17p) and del(11q), thus contributing to HGC (16–19). CLL cells demonstrate telomere dysfunction-induced foci (TIF) (20), marked by gamma H2AX and 53BP1 localization, and exhibit telomere deletions and terminal duplications (21). Short TL correlates with poor prognostic deletions (17p, 11q) and inferior survival (22–24). The relationship between TL and IGHV mutation status is established, with shorter telomeres in U-CLL cases, and a significant positive correlation between TL and mutational load at the IGHV locus (17, 25, 26). TL in mutated IGHV CLLs (M-CLL) is more heterogeneous, perhaps the result of variable numbers of cell division in response to antigen-mediated germinal centre reactions, where telomerase is activated to maintain TL (26–28), Analysis of the CLL methylome identified three clinically relevant epitypes linked to CpG modifications in naive (n-CLL) and memory (m-CLL) B cells, along with a third epitype with intermediate (i-CLL) methylation and moderate IGHV mutations, particularly enriched for IGLV3–21 R110 and SF3B1 mutations (29–31). These epitypes correlate with clinical outcomes (32) and differ in frequencies of TP53 aberrations and TL (33).

Our hypothesis was that the inclusion of these additional biomarkers, with GC, into clinical models of patient outcome will refine and improve the detection of patients destined to exhibit aggressive or progressive disease, with inferior overall survival. Our study was the first to evaluate the clinical relevance and independent prognostic significance of GC, TL, IGHV status, and methylation epitypes in 495 untreated CLL patients in UK (immuno)-chemotherapy trials. We demonstrated that, in addition to GC, epigenetic information and TL data can help identify additional patients with poor survival.

## Methods

### Patient Cohort

Our cohort comprised a total of 495 treatment-naïve CLL patients enrolled in three randomized clinical trials; the UK LRF CLL4 trial (34) (NCT00004218, n = 251); UKCRN ADMIRE (35) (‘ADM’, ID6897, n = 122); UKCRN ARCTIC trial (‘ARC’, ID7136, n = 122) (36) (Table S1). All patients were diagnosed using established morphological and immunophenotypic criteria. Informed consent was obtained from all participants in accordance with the Declaration of Helsinki, and the study was approved by the Somerset Regional Ethics Committee. The assessment of established biomarkers including FISH, CD38, ZAP70, and IGHV mutational status was performed at randomization as previously described (34–36). The distribution of clinico-biological features was consistent across cohorts (Table S2).

### Targeted Sequencing

Mutational data previously generated using an Illumina TruSeq Custom Amplicon panel was available for 454 patients (CLL4 n=228 (37) and ARC/ADM n=226 (38)). An additional 41 patients (CLL4 n=23, ARC/ADM n=18) were analysed using a bespoke Agilent Sureselect XT HS2 Targeted Enrichment System, targeting 63 genes selected for their clinical relevance in b-cell malignancies, according to manufacturer’s protocols. Mutations were identified using custom pipelines and filtered as previously reported (37) (Supp Methods). Consensus mutational data was available for 9 recurrently mutated CLL genes (Table S3).

### Copy number profiling

Previously published copy number alteration (CNA) data derived from Affymetrix SNP 6.0 (CLL4, n=108) (39, 40) and the HumanOmni2.5-8.0 SNP (ARC/ADM, n=212) (38) were available. An additional 32 ARC/ADM patients were profiled as previously described (32), using the Illumina Infinium Human Methylation 450 BeadChip (Illumina, Hayward, CA, USA), according to manufacturer’s instructions, at the Genomics and Proteomics Core Facility of the DKFZ (Heidelberg, Germany). Data processing was performed using RnBeads v2.93 (RRID:SCR_010958) and Conumee (41) (Supp Methods). The remaining 143 CLL4 patients were profiled using shallow WGS, using the Agilent SureSelect QXT system according to manufacturer’s recommendations as previously described (37, 42). The data was analysed using custom pipelines (Supp Methods).

After passing the inclusion criteria for individual technologies (Supp Methods), all CNA were manually curated by two independent experienced researchers, with 1295 CNA, ranging from 7Kb-155MB passing inspection. A high concordance for calling del17p, del11q, Tri12 and del13q was found between the four genomic technologies and FISH data (percentage agreement; 71-99%) (Table S4). Based on CNA burden, including aneuploidy, patients were classified as having Low Genomic Complexity (LGC, 0–2 CNAs, n=334/67%), Intermediate GC (IGC, 3–4 CNAs, n=97/20%) or High GC (HGC, ≥5 CNAs, n=64/13%). CNA identified within known regions of importance (2p, 4p, 6q, 11q, 13q, 14q and 17p) were included in all analyses, regardless of size, while a cutoff of ≥5 Mb was applied for other CNAs, as previously reported (43). Copy-number neutral loss of heterozygosity and biallelic losses, were excluded and counted as two CNAs, respectively. CNAs within ≤5 Mb of each other and putative chromothripsis events were counted as a single event.

No significant difference was observed in the proportion of each GC group in the CLL4 and ARC/ADM cohorts (Fig S1). The prevalence of CNAs across the four platforms was also highly concordance, except for a modest reduction in the CNAs detected by sWGS (Fig S2), due to more cautious CNA definition consequent on higher, but acceptable levels of background noise. To emphasize this point further, the sWGS data demonstrated a high concordance with FISH for del17p (98%), del11q (91%), Tri12 (95%) and del13q (80%) (Table S4).

### Telomere length analysis

Telomere length data was generated using Single Telomere Length Analysis (STELA) (ARC/ADM n=232) and monochrome multiplex PCR (MMQPCR) (CLL4 n=180) as previously described (22, 24). For 80 CLL4 patients analysed with both MMQPCR and STELA, a significant correlation between technologies was observed (Kendall’s rank correlation; 0.657 correlation coefficient). Additional profiling, using MMQ-PCR, was performed on 71 CLL4 and 12 ARC/ADM patients and statistical analyses were performed on the combined TL dataset (n=495), as previously reported (22).

### DNA Methylation Analysis

Previously published DNA methylation data was available on 446 patients (CLL4=226, ARC/ADM=220), derived from pyrosequencing (32). Patients were classified as naive B-cell–like CLL (n-CLL), memory B-cell–like CLL (m-CLL), and intermediate CLL (i-CLL), using established criteria (32, 44).

### Statistics

Clinico-biological associations were determined using Wilcoxon rank sum, Fisher’s exact and chi squared tests (significance; p=0.05), with Benjamini-Hochberg correction for multiple testing. Overall survival (OS) was measured in months from date of randomization to date of death from any cause or date of last follow-up, and progression free survival (PFS) was measured in months from date of randomization to progression (relapse requiring further treatment), date of death or date of last follow-up. CLL4 survival data (n=251) for PFS and OS came from the final updates in 2010 (10 years follow up) and 2016 (17 years follow-up), respectively (median PFS =2.4 years, median OS=6 years). ARC/ADM PFS and OS data (n=244) came from the 2022 update, after 9 years follow up (median PFS = 4.72 years, median OS=6.44 years). Kaplan-Meier curves (log-rank test), Cox proportional hazards models and Multivariate Cox Proportional Hazard models using stepwise backwards elimination were used to investigate PFS and OS. Analyses were performed in R(v4.3.0), STATA (v12.1) and SPSS (v23).

## Results

### Cohort overview

The overview of our cohort design and methodological approaches is depicted in Fig 1A. Baseline clinico-biological characteristics of the 495 CLL patients are shown in Table S2. Of these, 367 (74%) were male and 128 (26%) were female, with a median age of diagnosis of 64 years (range: 36-86 years). IGHV mutational status was available for 439 patients, of whom 39% (n=169) had mutated IGHV (M-CLL) and 60% (n=270) had unmutated IGHV (U-CLL) (Fig 1B). Telomere length ranged from 1.13-10.24Kb (median 3.21kb), with patients classified as TL-Short (<2.921lkb, n=193), TL-Intermediate (2.92–3.571lkb, n=123) or TL-Long (>3.571lkb, n=179) using established cut-offs (22) (Fig 1B). Additionally, methylation-based classification, according to published criteria (44), assigned patients to; n-CLL (n=218, 49%), i-CLL (n=144, 32%) or m-CLL (n=84, 19%) subgroup (Fig 1B).

**Figure 1.**
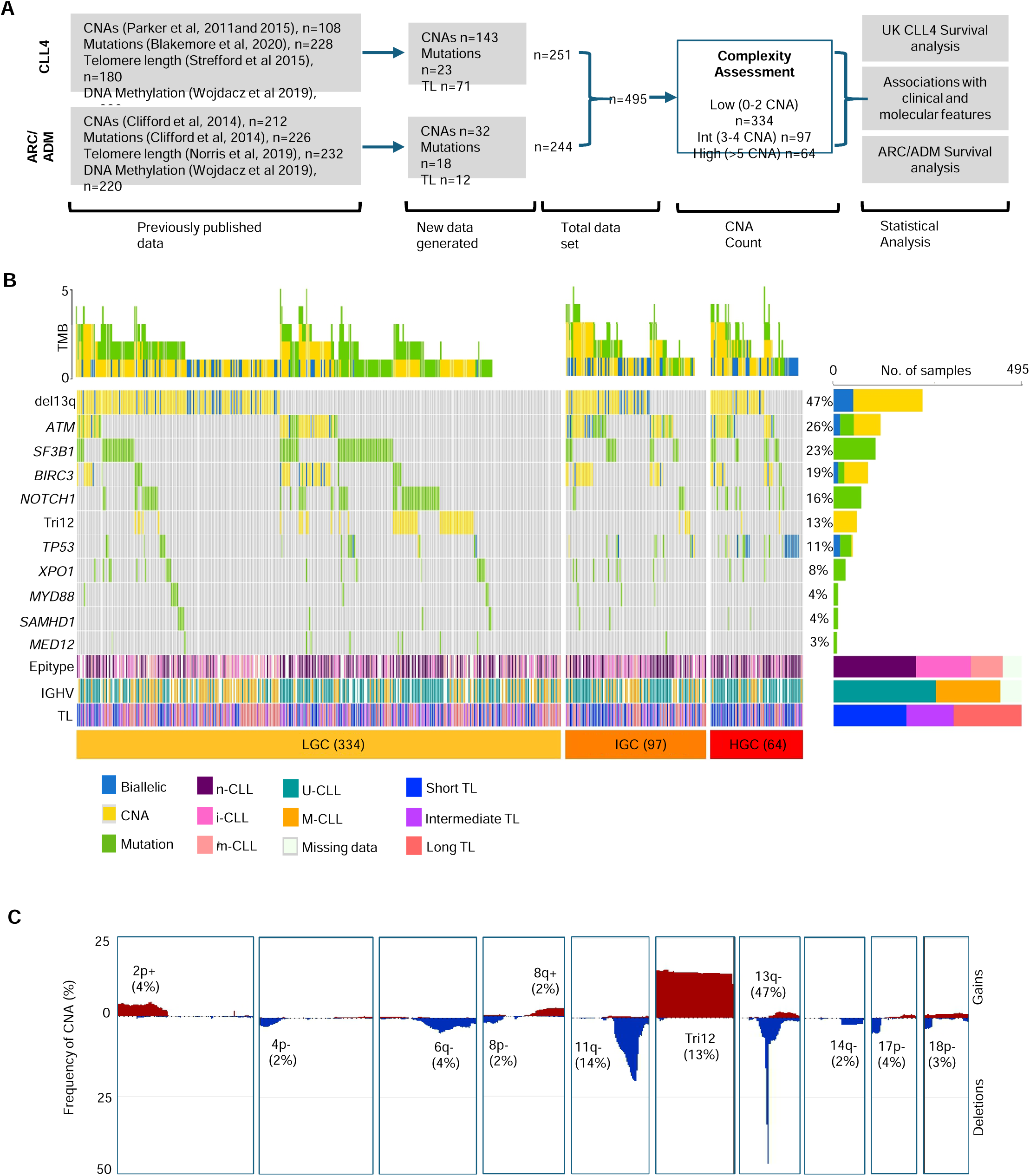
The genomic landscape of CLL A Consort diagram showing the experimental workflow and key datasets in our study. B Waterfall plot, segmented by genomic complexity classification (LGC, IGC and HGC), showing the frequency of mutations in the nine profiled genes (green bars) and most common CNA (yellow bars) in the total CLL cohort (n=495). Biallelic loss (deletion + mutation, or biallelic deletion = blue bars). Horizontal bars below show the distribution of epitype groups, IGHV usage, and telomere length (TL) for each patient, across the GC subgroups. C Frequency plot showing the percentage of patients with recurrent copy-number gains (red) and losses (blue) in 495 CLL patients.

In our cohort, 426 variants were identified in 284 patients across the nine consensus genes (ATM, SF3B1, NOTCH1, TP53, BIRC3, XPO1, MYD88, SAMHD1 and MED12), with a median of 0.86 (range: 0-6) mutations/sample. Mutations were detected in all genes, with the highest frequencies identified in SF3B1 (22%, n=111), NOTCH1 (15%, n=74), ATM (11%, n=54), TP53 (9%, n=47) and BIRC3 (6%, n=28) (Fig 1B).

Copy number analysis identified 1297 CNAs in 452/495 patients (mean: 2.62, range: 0-23), including 65 aneuploidy events (Tri12 n=62). Established CLL-associated CNA were identified at expected frequencies; del17p in 4.4%, del11q in 18%, del13q in 47.4%, tri12 in 12.5% (Fig 1B-C). Additional recurrent CNA (present in >2% of the cohort) and minimally deleted or enhanced regions were defined on chromosomes 2p, 4p, 6q, 8 and 14 (Fig 1C, Table S5).

### Genomic Complexity associates with clinico-biological features

First, we analysed the relationship between genomic complexity (GC) subgroups and key clinico-biological features of CLL. HGC was more prevalent in U-CLL (81%, n = 47, p < 0.001) (Fig 2A), with significantly higher CNA counts in U-CLL (mean: 2.86) compared to M-CLL (mean: 2.01, p<0.001). TL varied significantly across GC groups, with TL-L and TL-S predominantly occurring in LGC (41%), and HGC patients (61%), respectively (p<0.05) (Fig 2Bi). As a continuous variable, TL negatively correlated with CNA count (Kendall’s τ =-0.147, p < 0.001) (Fig 2Bii).

**Figure 2.**
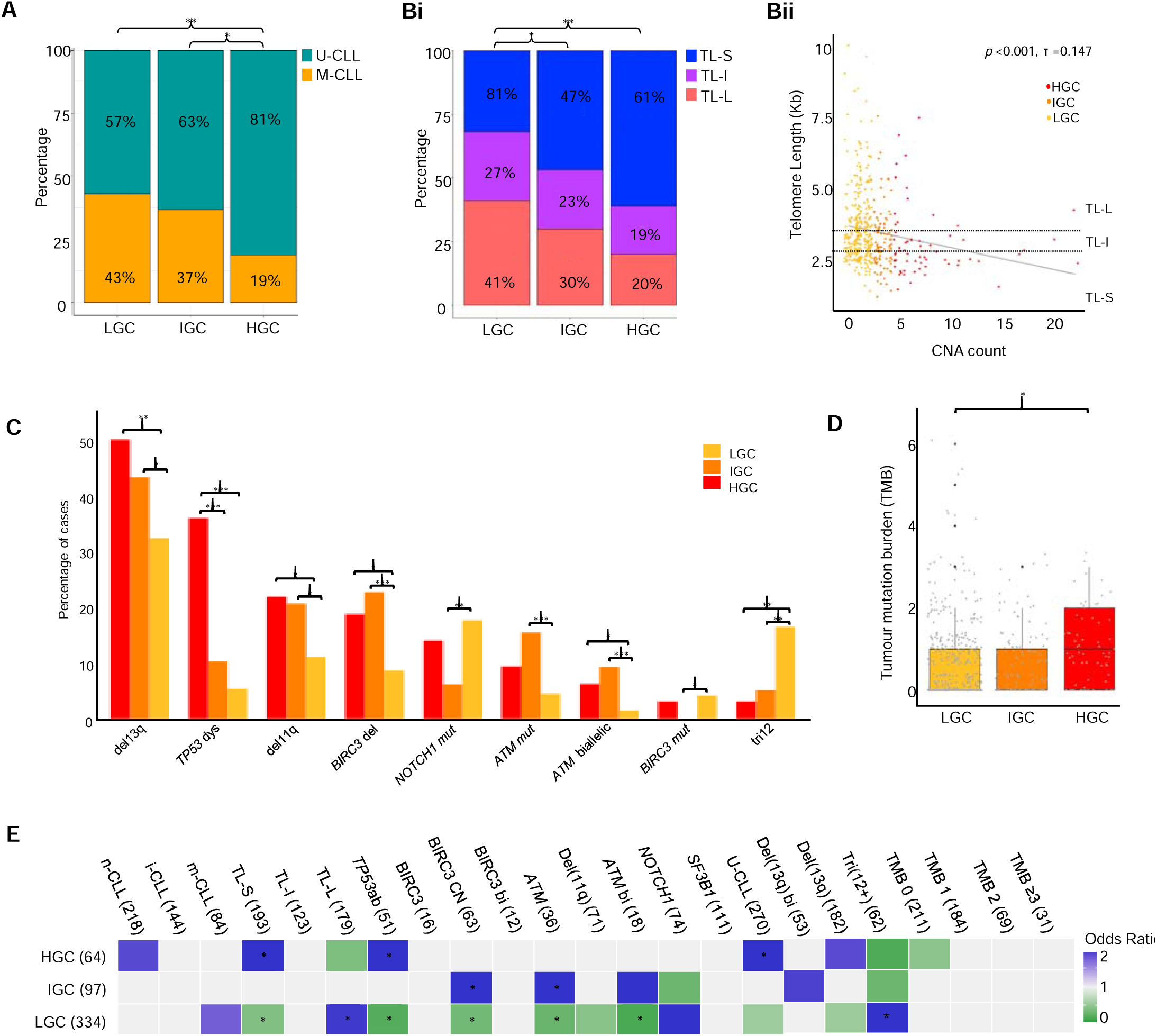
Genomic complexity associates with clinico-biological features A Stacked bar graph showing the proportion of U-CLL (teal) and M-CLL (orange) patients across the 3 GC subgroups. Bi Stacked bar graph showing the proportion of patients classified as having TL-S (blue), TL-I (purple) and TL-L (pink) across the 3 GC subgroups. Bii Scatterplot showing the significant negative correlation (p<0.001) between telomere length and CNA count. The linear regression model is depicted by the grey line. Points are coloured to indicate GC subgroup (HGC =red, IGC=orange, LGC=yellow). Dotted lines indicate the size cut-offs (Kb) for TL-S, TL-I and TL-L. C Frequency bar chart showing the features that are significantly associated with HGC, IGC and LGC (red, orange and yellow, respectively). Significant enrichment of features was determined using a Fishers Exact test. D Box and whisker plots showing the increased tumour mutation burden (TMB) in HGC patients compared to LGC and IGC. In all plots * indicates p<0.05, ** indicates p<0.001. E Association plot of odds ratios showing features that co-occur with GC subgroups, or are mutually exclusive (blue and green squares, respectively) (p≤0.05, corrected p-values calculated using Benjamini-Hochberg FDR (p≤0.05 *).

GC was associated with recurrent chromosomal abnormalities; Monoallelic deletions of chromosome 13q were significantly enriched in HGC (n=32, 50%) compared to LCG (n=108, 32%, p<0.01) (Fig2C). As 13q deletion size has previously been linked to GC, we further classified our deletions into Class I (<2Mb, n=86) and Class II (>2Mb, n=170) deletions (39), with Class II deletions significantly enriched in HGC (45%) and IGC (49%) (p<0.05). Trisomy 12 was more prevalent in LGC (n=55, 16.5%) versus IGC (n=5, 5.2%) and HGC groups (n=2, 3.1%) (p<0.01) and frequently co-occurred with NOTCH1 mutations in 24/62 cases, with 22 (92%) occurring in LGC (Fig2C).

Deletions of 11q, encompassing the ATM gene were detected in 71 patients, and were enriched in HGC and IGC (n=14 [22%] and n=20 [21%], respectively). Additionally, 54 patients harboured either a mutation (n=36), or a deletion and a mutation of ATM (n=18), both of which were significantly enriched in IGC (p<0.001) (Fig2C). BIRC3 deletions (n=63) did not occur independently of ATM loss and were also significantly associated with HGC and IGC cases (n=18 and n=22, respectively, p<0.001), with no significant associations recorded between GC groups and biallelic BIRC3 inactivation (n=12) (Fig2C).

TP53 dysfunction, defined as copy number loss, mutation or both, was observed in 51 patients, with 23 (36%) classified as HGC (36%) (p<0.001) (Fig2C). Notably, 14 of the 18 cases (78%) with biallelic TP53 loss were HGC (Fig1B). A modest positive correlation was observed between Tumour Mutation Burden (TMB) and CNA count (Kendall’s τ=0.26, p=0.042), with a higher mean mutation count in the HGC compared to LGC (mean= 1.08 vs 0.83, respectively, p<0.05) (Fig 2D). NOTCH1 (n=74) and BIRC3 mutations (n=16) were more frequently observed in LGC (17 and 8%, respectively), and whilst SF3B1 mutations were not overrepresented in any complexity subgroup, 70% of SF3B1 mutations (n=79) were found in LGC patients. While GC was not significantly associated with methylation-based subgroups, 62% (n = 38) of HGC patients belonged to the n-CLL subgroup (Fig 2E).

### The clinical implications of High Genomic Complexity

Due to the different treatment modalities and follow up periods for ARC/ADM and CLL4, survival analysis was performed separately for each cohort. In both cohorts, HGC was significantly associated with a shorter median PFS and OS compared to LGC (Table S6), as well as significantly lower 5-year survival rates (ARC/ADM; 51% vs 29%, CLL4; 61% vs 42%) (p<0.01). Univariate Cox regression (UCR) analysis of 13 clinical and molecular features was performed to evaluate their prognostic significance for PFS and OS (Tables S7-8), with significant associations displayed in Fig 3Ai and Bi.

**Figure 3.**
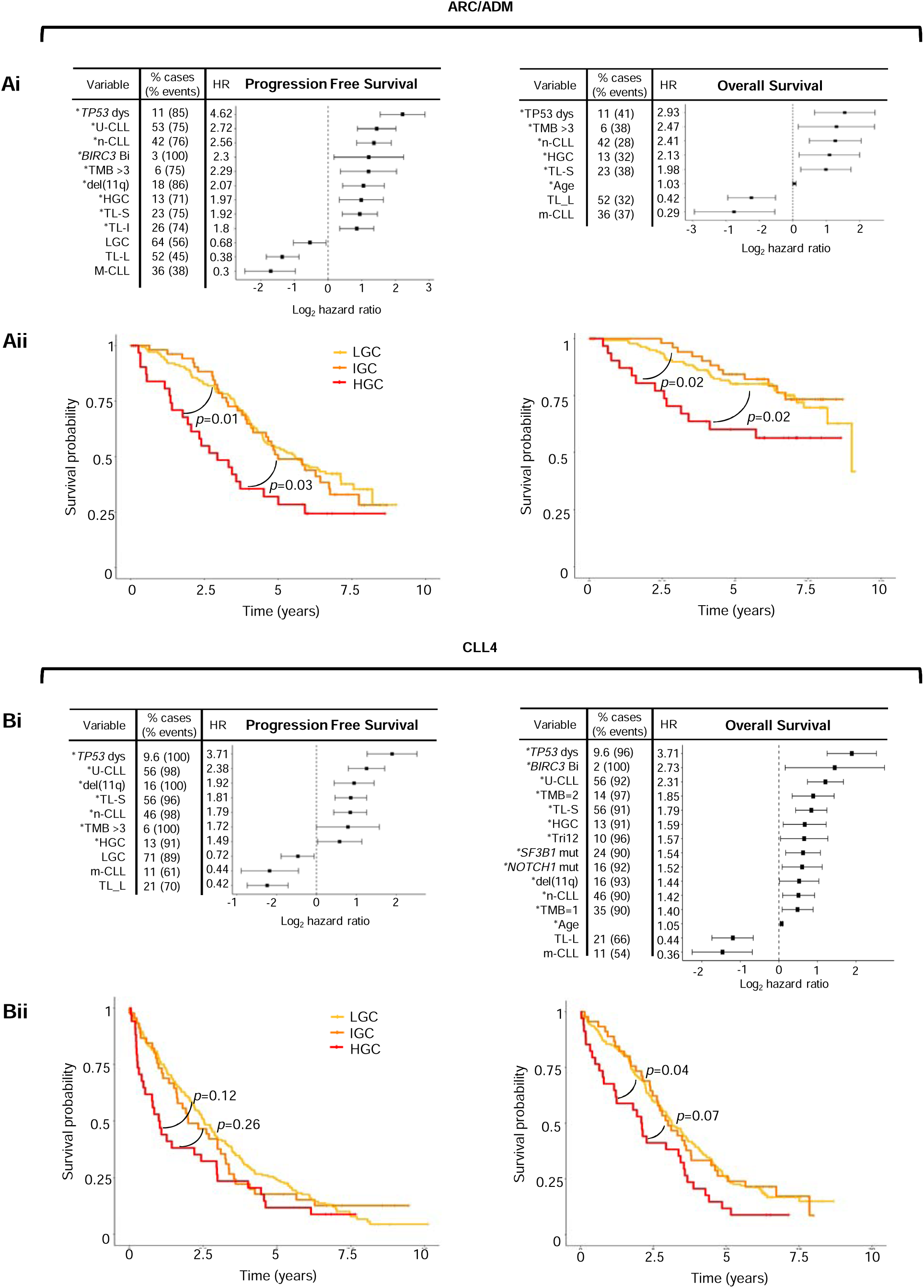
Clinical implications of GC in univariate analysis Ai-Bi Forest plots showing the hazard ratios for variables significant (p<0.05) for Progression Free Survival (PFS) and overall survival (OS) in univariate cox regression (UCR) analysis, for ARC/ADM (Ai) and CLL4 (Aii). The proportion of cases positive for each variable, and the proportion of events in those cases is shown. * indicates poor risk variables included in the multivariate model (MV). Aii-Bii Kaplan-Meyer plots showing the shorter PFS and OS in HGC cases in ARC/ADM (Aii)and CLL4 (Bii).

In both cohorts, HGC was associated with shorter PFS (ARC/ADM: HR=1.97, p=0.002; CLL4: HR=1.49, p=0.04) and OS (ARC/ADM: HR=2.13, p=0.015; CLL4: HR=1.59, p=0.018). Additionally, several established poor prognostic indicators for PFS were identified in both cohorts including TP53 dysfunction, n-CLL classification, short or intermediate TL, U-CLL and del(11q). TMB>3 mutations, also emerged as a significant predictor of shorter PFS in ARC/ADM and CLL4 (ARC/ADM: HR=2.29, p=0.003; CLL4: HR=1.72, p=0.046). Reduced OS in the ARC/ADM cohort was predicted by age, short or intermediate TL, TP53 dysfunction, n-CLL classification, and TMB>3 (Fig 3Ai). In the CLL4 cohort, in addition to these variables, a further seven features were associated with shorter OS, including NOTCH1 and SF3B1 mutations (Fig 3Bi). Additionally, Kaplan-Meier analysis of HGC patients versus IGC and LGC patients, confirmed shorter PFS in ARC/ADM, and shorter OS in both ARC/ADM and CLL4 (Fig3Aii and Bii).

Given the co-occurrence of HGC with other poor-risk biological features, we next estimated the adjusted impact of HGC on PFS and OS by controlling for these confounders using multivariate Cox proportional hazard analysis with stepwise backward selection. We incorporated poor risk variables, identified through UCR analysis (Fig3Ai and Bi). For PFS, in both ARC/ADM (9 variables tested, 200 patients/118 events) and CLL4 (7 variables tested, 198 patients/176 events), HGC was not a significant independent predictor in the final model (Fig 4A). Instead, the models included TP53 dysfunction (ARC/ADM: HR=3.59, p<0.001; CLL4: HR=2.68, p=0.01), unmutated IGHV (ARC/ADM: HR=2.04, p=0.001; CLL4: HR=1.94, p<0.01) and TL-S (ARC/ADM: HR=1.92, p=0.005; CLL4: HR=1.52, p<0.01) (Fig 4A). In the ARC/ADM cohort, a model incorporating six high risk variables (220 patients/58 events), identified TP53 dysfunction (HR=2.91, p=0.002), n-CLL (HR=1.94, p=0.02) and age (HR=1.05, p=0.004) as independent predictors of shorter OS (Fig 4A). In the CLL4 cohort, a multivariate model for OS, which included 12 poor risk variables, and age, (198 patients/167 events), resulted in a final model with seven independent predictors of shorter OS. These included HGC (HR=1.61, p=0.02), TP53 dysfunction (HR=2.94, p<0.001), Tri12 (HR=1.79, p=0.002), TL-S (HR=1.7, p=0.002), U-CLL (HR=1.54, p=0.02), SF3B1 mutations (HR=1.5, p=0.02) and age (HR=1.05, p<0.001) (Fig 4A).

**Figure 4.**
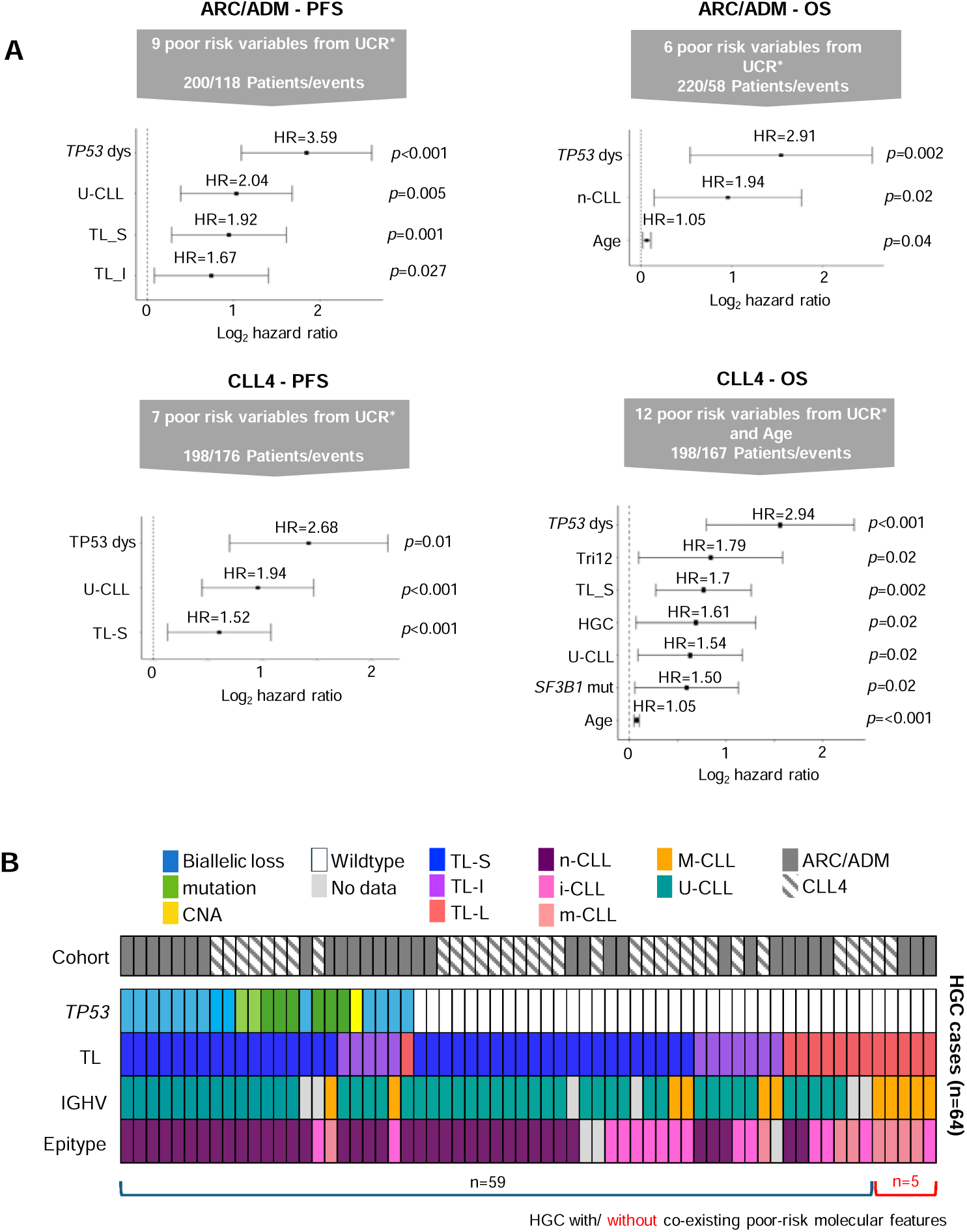
Clinical implications of GC in multivariate modelling A Forest plots showing the hazard ratios for variables significant (p<0.05) for Progression Free Survival (PFS) and overall survival (OS) in multivariate (MV) analysis, for ARC/ADM and CLL4. Grey arrows describe the number of poor risk variables and the number of patients and events included in each MV model. Forest plots show the significant independent predictors of outcome in each MV model. B Plot showing the distribution of high-risk features in the 64 HGC cases.

Multivariate modelling did not consistently identify HGC as an independent predictor of shorter PFS or OS, suggesting that it may act as a surrogate for other high-risk features, which may underlie the observed complexity. Focusing on variables highlighted in our multivariate analysis, we examined the 64 HGC cases across our combined cohorts. Of these, 23 patients exhibited TP53 dysfunction, of which 14 also showed co-occurrence of U-CLL, TL-S, and n-CLL; 5 had U-CLL and n-CLL; and 3 had TL-S. Among the remaining 41 TP53 wild-type HGC cases, all but five (7.8%) harboured at least one additional high-risk feature: TL-S (n=18), U-CLL (n=26), or n-CLL (n=17) (Fig. 4B). Taken together, our association and survival analyses show that while HGC is linked to poor survival in univariate models, its prognostic value is attenuated in multivariate analysis. Features such as TP53 dysfunction, unmutated IGHV, short telomere length, and methylation subtype are strongly associated with HGC and independently predict outcome, suggesting that these biomarkers, individually or in combination, may more precisely define this high-risk patient subgroup.

## Discussion

This comprehensive (epi)genomic analysis of GC in a large cohort of 495 clinical trial patients underscores the intricate interplay between GC and other high-risk biological features, as well as their collective impact on clinical outcomes. We confirm and extend previous findings by elucidating the relationship between GC and other prognostically relevant molecular characteristics, such as the enrichment of TP53 lesions and unmutated CLL (U-CLL) in HGC cases (3–5, 8), along with an increased prevalence of trisomy 12 in LGC cases (1).

Notably, our work is the first to include contemporary cell-of-origin measures to identify n-CLL cases and to assess TL, which is a consequence of cellular proliferation. We hypothesized that these measures would better define poor-risk patients not fully captured by traditional GC metrics. Our findings support this, as TL and n-CLL independently identified poor-risk patients, with 92.2% of HGC cases showing TP53 aberrations, U-CLL, short TL, or being classified as n-CLL. While current clinical paradigms focus on GC, TP53, and U-CLL for prognostic purposes, our data suggest future research should emphasize DNA methylation signatures and telomere attrition for identifying patients at risk for aggressive disease.

Our work confirms the link between TP53 dysfunction, TL shortening, and HGC in CLL. Loss of TP53, via mutation or 17p deletion, impairs the DNA damage response, allowing genomic lesions to bypass checkpoints and continue dividing, leading to chromosomal instability (17, 21). ATM deletions were enriched in both HGC and IGC groups, while mutations were more common in IGC, indicating a complex relationship between ATM status and GC. Unlike CLL B-cells with ATM mutations, those with 11q deletion alone show normal levels of ATM S1981 auto-phosphorylation and p53 S15 phosphorylation after ionizing radiation (45). Our analyses did not show greater GC in bi-allelic ATM cases, as might be expected based on functional data, which is in keeping with previous TL data (46), suggesting that other 11q sequences like miR-34 and H2AFX may influence GC through altered DNA damage response (47–49). BIRC3 mutations were enriched in LGC CLL, reflecting its role as a negative regulator of NF-κB signalling rather than a direct role in DNA damage response (50). The size of 13q deletions correlates with genomic complexity and disease progression in CLL (51), with large deletions encompassing genes like RB1 and RNASEH2B. Loss of RNASEH2B impairs RNase H2 activity, sensitizing CLL cells to PARP inhibitors through defective ribonucleotide excision repair (52). In our study, 66.4% of 13q deletions were large (Class II) (39), suggesting a role in genomic instability and potential sensitivity to PARP inhibition. Mutations in NOTCH1 and SF3B1 displayed variable distributions across genomic complexity subgroups and did not independently stratify patient outcomes, with the co-occurrence of NOTCH1 mutations and trisomy 12 in LGC cases highlighting the heterogeneity of molecular interactions in CLL pathogenesis. A correlation between TL and GC suggests a potential causative link, with telomere shortening possibly preceding GC (16, 53). Transformation may stabilize TL in B-cells, promoting breakage-fusion-bridge cycles and genomic instability. Alternatively, GC may reflect a more proliferative disease, especially in U-CLL with enhanced BCR signalling (54). If TL shortening precedes GC, telomerase inhibition could disrupt this cycle; otherwise, it may impair the expansion of B-CLL-specific T cells in patients with short TL (55).

HGC was linked to PFS and OS in both the ARC/ADM and CLL4 cohorts, but it did not consistently maintain prognostic independence in multivariate models with TP53 abnormalities, TL, n-CLL, and IGHV status. This suggests that HGC may primarily act as a surrogate for high-risk biological alterations rather than an independent predictor of poor prognosis. Notably, nearly half of HGC cases harboured TP53 abnormalities. Biallelic TP53 loss, an indicator of therapeutic resistance, was especially common in the HGC subgroup, illustrating the role of genomic instability in clonal evolution and resistance mechanisms in CLL. Furthermore, most TP53 wild-type HGC cases exhibited at least one other high-risk feature (e.g., short-TL, U-CLL, or n-CLL epitype), reinforcing the idea that GC is rarely an isolated phenomenon but rather reflects converging pathways of adverse biology.

This study has several strengths, including: 1) a large, well-characterized cohort with extended clinical follow-up and independent validation in two clinical trials; 2) comprehensive data that build detailed biomarker profiles for each trial participant, including biological variables like TL and DNA methylation epitype, which have not been clinically assessed together with GC until now; 3) the chemotherapy backbone used in these trials, which may provide valuable insights into the association between GC and other biomarkers due to established links between these therapies and genomic instability; and 4) the ongoing and future relevance of chemotherapy with or without rituximab in resource-limited settings (56), underscoring the need to understand the biological drivers of poor responses to these agents.

However, several limitations must be acknowledged. Firstly, the trials assessed traditional (immuno)- chemotherapy regimens that now have limited front-line use in high-income health systems, which may restrict the applicability of our findings in the era of targeted therapies. However, our work accentuates the necessity to investigate the combination of biomarkers in trials of these agents, as sophisticated measures of cell-of-origin, cellular proliferation, and genomic instability will likely remain relevant. Secondly, a broader gene panel could enhance our understanding of the genomic mechanisms underlying GC, particularly regarding POT1 mutations, which lead to the accumulation of telomeric and chromosomal abnormalities (57). Additionally, our study utilized CNA profiling with limited resolution that cannot identify balanced translocations, which, while rare, represent a significant proportion of CLL translocations. The broader literature also demonstrates inconsistency in defining and assessing GC, with varying technologies and thresholds hindering the establishment of a standardized, clinically meaningful metric. Importantly, our study suggests that other biomarkers can effectively identify these poor-risk patients without requiring consensus on GC.

In conclusion, our data support a model in which HGC in CLL is a composite biomarker of adverse biology, closely associated with TP53 dysfunction, telomere erosion, and unmutated IGHV status, particularly those U-CLL with the most naïve epigenetic features. Future efforts should focus on longitudinal profiling to map the dynamics of complexity acquisition with telomere attrition in cases characterized at the epigenetic level, with integration of these observations into integrative risk models to guide precision management strategies in CLL.

## Data Availability

All data produced in the present study are available upon reasonable request to the authors

## Acknowledgements

The authors gratefully acknowledge all patients who contributed to this study. The authors are indebted to Professor Daniel Catovsky for the generation and curation of the CLL4 trial. This work was funded by Bloodwise (11052, 12036), the Kay Kendall Leukaemia Fund (873), Cancer Research UK (ECRIN-M3 accelerator award C42023/A29370, Southampton Experimental Cancer Medicine Centre grant C24563/A15581, Cancer Research UK Southampton Centre grant C34999/A18087, and programme C2750/A23669) and the Bournemouth Leukaemia Fund. The LRF CLL4 trial was funded by a core grant from Leukaemia and Lymphoma Research. Patient material was obtained from the UK CLL Trials Biobank, University of Liverpool, which is funded by Blood Cancer UK. ME acknowledges the support by The Arbib Charitable Fund. The views expressed in this paper are those of the authors and not necessarily those of the funding agencies. LC received a PhD studentship funded by Cancer Research UK and the Medical Research Council. The Baird lab is funded by Cancer Research UK programme C17199/A29202.

## Contributions

LC, HP, KN, ANT, HA performed the experimental work; AP performed the molecular diagnostic assays; HP, LC, LK, ME, DB and JG conducted the statistical and bioinformatics analyses; ME, AP, PH, AS, RW and DGO contributed patient samples and data; JCS and CP initiated and designed the study; HP, LC and JCS wrote the paper and all authors critically reviewed the final paper.

---

**Supplementary Figure 1.**
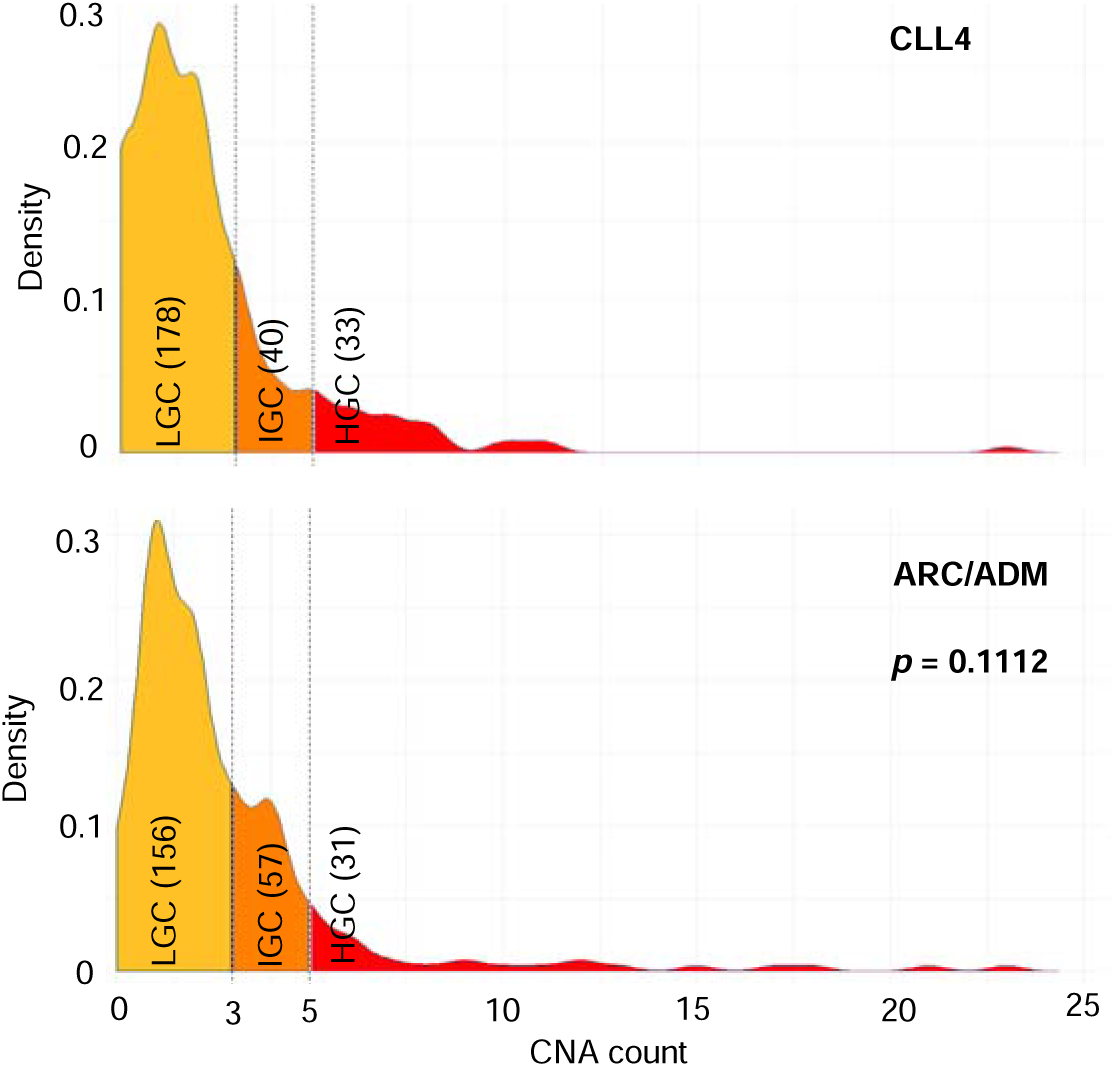
Copy number count and genomic complexity subgroup distribution across cohorts. Density plot showing the copy number counts and equal distribution of GC subgroups across the CLL4 and ARC/ADM cohort (top and bottom plots respectively) (Pearson’s Chi squared *p*=0.0112).

**Supplementary Figure 2.**
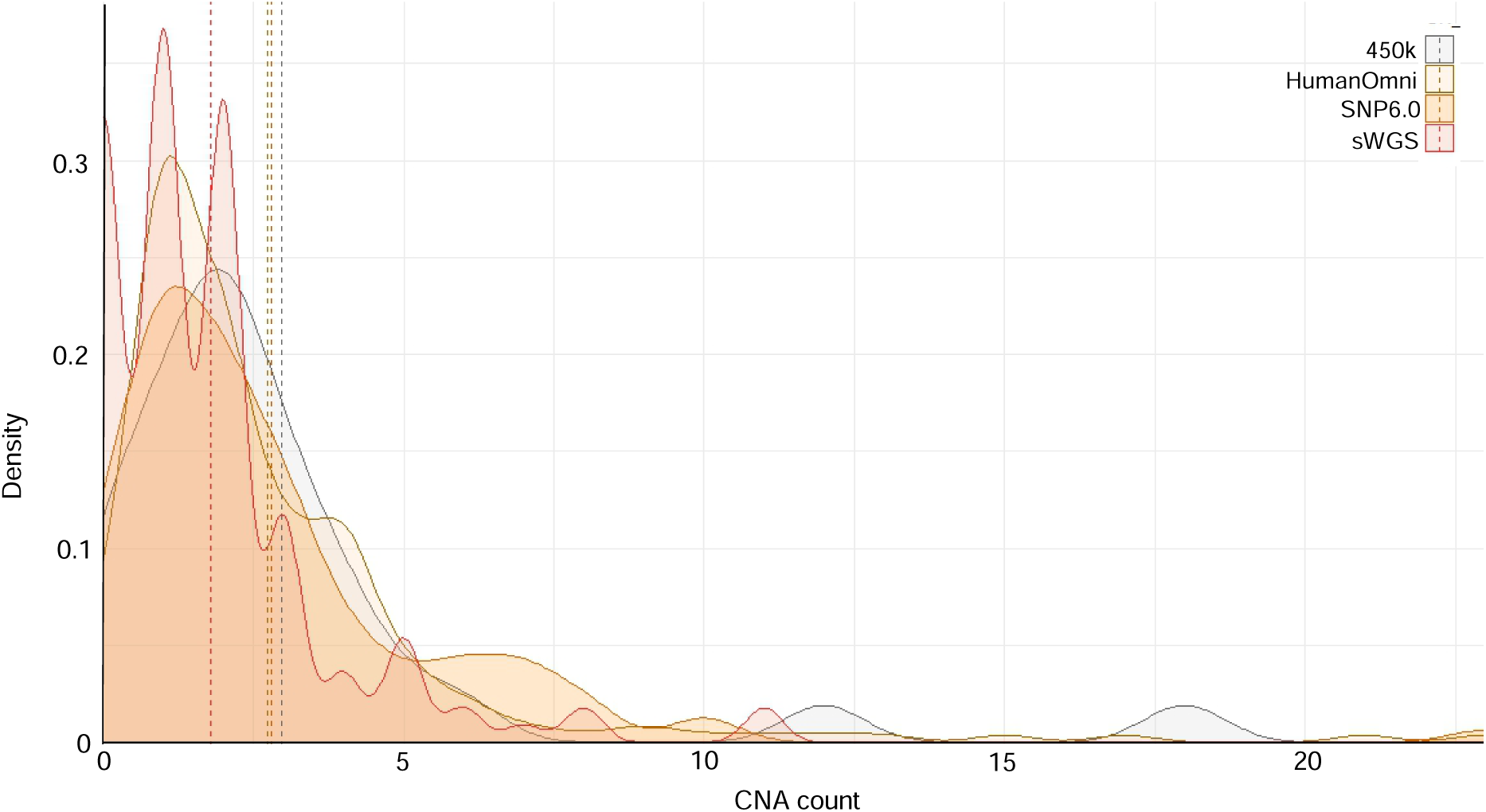
Distribution of CNA counts across sequencing technologies. Density plot showing the distribution of copy number counts across the 4 sequencing technologies

