## Supplementary Tables for "High-risk Molecular Features Eclipse Genomic Complexity in Predicting CLL Patient Outcomes; Insights from the UK CLL4, ARCTIC and ADMIRE Trials"

**Supplementary Materials**

Supplementary Table 1. Description of clinical trials used in the study

| **Clinical Trial** | **Phase** | **Treatment arms** | **Total**  **N (%)** | **In this study**  **N (%)** |
| --- | --- | --- | --- | --- |
| CLL4 | III |  | 777 | 251 |
|  |  | Chlorambucil | 387 (50) | 86 (34) |
|  |  | Fludarabine | 194 (25) | 108 (43) |
|  |  | Fludarabine + Cyclophosphamide | 196 (25) | 57 (23) |
| ARCTIC | II |  | 199 | 122^ |
|  |  | Fludarabine + Cyclophosphamide + Rituximab | 100 (50) | 56 (46) |
|  |  | Fludarabine + Cyclophosphamide + Mitoxantrone + miniRituximab | 99 (50) | 57* (47) |
| ADMIRE | II |  | 215 | 122^ |
|  |  | Fludarabine + Cyclophosphamide + Rituximab | 107 (50) | 54 (44) |
|  |  | Fludarabine + Cyclophosphamide + Mitoxantrone + Rituximab | 108 (50) | 59 (48) |

* includes 13 cases which started a FCMminiR regine but were crossed over to FCR. ^ 9 clinical trial patients have unknown treatment regime.

Supplementary Table 2. Baseline clinco-biological features of the ARCTIC, ADMIRE and CLL4 trial patients.

| **Variable** | **ARCTIC & ADMIRE**  **N (%)** | **CLL4**  **N (%)** | **Concordance**  **P-value** |
| --- | --- | --- | --- |
| Total number of patients | 244 | 251 |  |
| Age, median years (range) | 63 (36-80) | 64 (42-86) | 0.057 |
| Gender |  |  |  |
| Male | 183 (75%) | 184 (73.3%) | 0.67 |
| Female | 61 (25%) | 67 (26.7%) |  |
| Binet Stage |  |  |  |
| A | 35 (14.4%) | 64 (25.5%) | <0.01 |
| B | 127 (52%) | 110 (43.8%) |  |
| C | 82 (33.6%) | 77 (30.7%) |  |
| IGHV Mutation Status |  |  |  |
| U-CLL | 129/218 (59.2%) | 141/221 (63.8%) | 0.32 |
| M-CLL | 89/218 (40.8%) | 80/221 (36.2%) |  |
| TP53 Dysfunction |  |  |  |
| Wildtype | 217 (88.9%) | 227 (90.4%) | 0.58 |
| Present | 27 (11.1%) | 24 (9.6%) |  |
| ATM Disruption |  |  |  |
| Wildtype | 177 (72.6%) | 193 (76.9%) | <0.05 |
| Deleted | 30 (12.3%) | 41 (16.3%) |  |
| Mutated | 24 (9.8%) | 12 (4.8%) |  |
| Biallelic | 13 (5.3%) | 5 (2%) |  |
| BIRC3 Disruption |  |  |  |
| Wildtype | 201 (82.4%) | 203 (80.9%) | 0.62 |
| Deleted | 27 (11.1%) | 36 (14.3%) |  |
| Mutated | 9 (3.7%) | 7 (2.8%) |  |
| Biallelic | 7 (2.9%) | 5 (2%) |  |
| Trisomy 12 |  |  |  |
| Wildtype | 207 (84.8%) | 226 (90%) | 0.08 |
| Present | 37 (15.2%) | 25 (10%) |  |
| Del13q |  |  |  |
| Wildtype | 115 (47.1%) | 145 (57.8%) | 0.05 |
| Deletion | 104 (42.6%) | 81 (32.3%) |  |
| Biallelic deletion | 25 (10.2%) | 25 (10%) |  |
| SF3B1 |  |  |  |
| Wildtype | 193 (79.1%) | 191 (76.1%) | 0.71 |
| Mutated | 51 (20.9%) | 60 (23.9%) |  |
| NOTCH1 |  |  |  |
| Wildtype | 209 (85.7%) | 212 (84.5%) | 0.71 |
| Mutated | 35 (14.3%) | 39 (15.5%) |  |
| Epigenetic Subgroup |  |  |  |
| n-CLL | 103/220 (46.8%) | 115/226 (50.9%) | <0.01 |
| i-CLL | 61/220 (27.7%) | 83/226 (36.7%) |  |
| m-CLL | 56/220 (25.5%) | 28/226 (12.4%) |  |
| Telomere Length |  |  |  |
| Short | 56 (23%) | 137 (54.6%) | <0.01 |
| Intermediate | 62 (25.4%) | 61 (24.3%) |  |
| Long | 126 (51.6%) | 53 (21.1%) |  |
| Telomere Length, median kb (range) | 3.63 (1.13-7.68) | 2.88 (1.93-10.24) | <0.01 |
| CNA count, median (range) | 2 (0-26) | 2 (0-23) | 0.01 |
| Mutation Count |  |  |  |
| 0 | 95 (38.9%) | 116 (46.2%) | 0.42 |
| 1 | 97 (39.8%) | 87 (34.7%) |  |
| 2 | 35 (14.3%) | 34 (13.5%) |  |
| ≥3 | 17 (7%) | 14 (5.6%) |  |
| Genomic Complexity |  |  |  |
| Low | 156 (63.9%) | 178 (70.9%) | 0.11 |
| Intermediate | 57 (23.4%) | 40 (15.9%) |  |
| High | 31 (12.7%) | 33 (13.1%) |  |

CNA data are generated from genomic techniques, not FISH. Chi squared was used to test the concordance between categorical variables and a Wilcoxon rank test was used for continuous variables.

Supplementary Table 3. Targeted Sequencing Panel designs and consensus genes

| Agilent HS2 Custom Panel (n=63) | CLL4 TruSeq Custom Panel (n=22) | ARC/ADM TruSeq Custom Panel (n=9) | Consensus genes |
| --- | --- | --- | --- |
| *ARID1A* | *ATM* | *ATM* | *ATM* |
| *ATM* | *BIRC3* | *BIRC3* | *BIRC3* |
| *BCL2* | *BRAF* | *MED12* | *MED12* |
| *BCOR* | *CHD2* | *MYD88* | *MYD88* |
| *BIRC3* | *DDX3X* | *NOTCH1* | *NOTCH1* |
| *BRAF* | *ERG2* | *SAMHD1* | *SAMHD1* |
| *BTK* | *FBXW7* | *SF3B1* | *SF3B1* |
| *CARD11* | *HIST1H1E* | *TP53* | *TP53* |
| *CCND3* | *KRAS* | *XPO1* | *XPO1* |
| *CD79A* | *MED12* |  |  |
| *CD79B* | *MGA* |  |  |
| *CDH23* | *MYD88* |  |  |
| *CHD2* | *NFKB1E* |  |  |
| *CHD2* | *NOTCH1* |  |  |
| *CREBBP* | *NRAS* |  |  |
| *CXCR4* | *POT1* |  |  |
| *DDX3X* | *RPS15* |  |  |
| *ERG2* | *SAMHD1* |  |  |
| *EZH2* | *SETD2* |  |  |
| *FBXW7* | *SF3B1* |  |  |
| *FLNC* | *TP53* |  |  |
| *HIST1H1E* | *XPO1* |  |  |
| *ID3* |  |  |  |
| *IDH2EX4* |  |  |  |
| *JAK3* |  |  |  |
| *KDM2B* |  |  |  |
| *KLF2* |  |  |  |
| *KLHL6* |  |  |  |
| *KMT2D* |  |  |  |
| *KRAS* |  |  |  |
| *LRP1B* |  |  |  |
| *LTB* |  |  |  |
| *MAP2K1* |  |  |  |
| *MAP3K14* |  |  |  |
| *MDM2* |  |  |  |
| *MED12* |  |  |  |
| *MGA* |  |  |  |
| *MYD88* |  |  |  |
| *NFKB1E* |  |  |  |
| *NOTCH1* |  |  |  |
| *NOTCH2* |  |  |  |
| *NRAS* |  |  |  |
| *PAX5 (NONCODING)* |  |  |  |
| *PLCG2* |  |  |  |
| *PLS3* |  |  |  |
| *POT1* |  |  |  |
| *PRKDC* |  |  |  |
| *PTPRD* |  |  |  |
| *RHOA* |  |  |  |
| *RPS15* |  |  |  |
| *SAMHD1* |  |  |  |
| *SETD2* |  |  |  |
| *SF3B1* |  |  |  |
| *SPEN* |  |  |  |
| *STAT3* |  |  |  |
| *TCF3* |  |  |  |
| *TET2* |  |  |  |
| *TNFAIP3* |  |  |  |
| *TNFRSF14* |  |  |  |
| *TP53* |  |  |  |
| *TRAF3* |  |  |  |
| *UBR5* |  |  |  |
| *USP34* |  |  |  |
| *XPO1* |  |  |  |

|  |  | **SNP 6.0 Array** | | **Cohen’s Kappa** | **sWGS** | | **Cohen’s Kappa** | **HumanOmni Array** | | **Cohen’s Kappa** | **450k Array** | | **Cohen’s Kappa** |
| --- | --- | --- | --- | --- | --- | --- | --- | --- | --- | --- | --- | --- | --- |
| FISH | Del17p |  |  |  |  |  |  |  |  |  |  |  |  |
|  |  | Yes | No |  | Yes | No |  | Yes | No |  | Yes | No |  |
|  | Yes | 5 | 1 | 0.82 | 5 | 2 | 0.83 | 8 | 5 | 0.71 | 0 | 1 | -0.045 |
|  | No | 1 | 94 | **98%** | 0 | 123 | **98%** | 1 | 191 | **97%** | 2 | 28 | **90%** |
|  | Del11q |  |  |  |  |  |  |  |  |  |  |  |  |
|  |  | Yes | No |  | Yes | No |  | Yes | No |  | Yes | No |  |
|  | Yes | 28 | 7 | 0.84 | 12 | 8 | 0.61 | 31 | 7 | 0.77 | 5 | 4 | 0.64 |
|  | No | 0 | 67 | **93%** | 4 | 107 | **91%** | 7 | 162 | **93%** | 0 | 22 | **87%** |
|  | Trisomy 12 |  |  |  |  |  |  |  |  |  |  |  |  |
|  |  | Yes | No |  | Yes | No |  | Yes | No |  | Yes | No |  |
|  | Yes | 9 | 2 | 0.84 | 13 | 5 | 0.79 | 5 | 1 | 0.90 | 4 | 1 | 0.85 |
|  | No | 1 | 90 | **97%** | 1 | 112 | **95%** | 0 | 74 | **99%** | 0 | 12 | **94%** |
|  | Del13q |  |  |  |  |  |  |  |  |  |  |  |  |
|  |  | Yes | No |  | Yes | No |  | Yes | No |  | Yes | No |  |
|  | Yes | 30 | 27 | 0.44 | 59 | 18 | 0.60 | 48 | 7 | 0.73 | 5 | 3 | 0.63 |
|  | No | 3 | 42 | **71%** | 8 | 46 | **80%** | 3 | 23 | **88%** | 0 | 8 | **81%** |

Supplementary Table 4. Concordance between FISH and copy number calling by genomic technologies.

Cohen’s kappa value is used to interpret the strength of the agreement between methods; <0=Poor, 0.01-0.2=Slight, 0.21-0.4=Fair, 0.41-0.6=Moderate, 0.61-0.8=Substantial and 0.81-1=Almost perfect. * A negative value indicates that the two technologies tend to disagree more then would be expected by chance. Percentage agreement is shown in bold.

Supplementary Table 5. Minimally deleted or enhanced regions (MDRs/MERs) observed in at least 2% (n=10) of patients.

| **Chromosome region** | **Frequency % (n=)** | **Start Genomic position (Mb)** | **End Genomic position (Mb)** | **Size (Mb)** | **No. Genes** | **Gene content** |
| --- | --- | --- | --- | --- | --- | --- |
| del13q | 39 (170^(23)^) | 49.97 | 50.13 | 0.16 | 4 | *DLEU1, TRIM13, KCNRG, DLEU2 (Mir16-1/Mir15A)* |
| del11q | 18 (89) | 108.21 | 108.31 | 0.10 | 3 | Includes *ATM* |
| del17p | 4.4 (22) | 7.56 | 7.84 | 0.28 | 14 | Includes *TP53* |
| del6q | 4.4 (22) | 107.43 | 109.44 | 2.01 | 14 | Many |
| gain2p | 4 (20) | 60.22 | 62.03 | 1.80 | 9 | Includes *REL, BCL11A, XPO1* |
| del18p | 3.2 (16) | 1.75 | 6.24 | 4.49 | 15 | Many |
| del4p | 2.6 (12^(1)^) | 7.12 | 15.99 | 8.87 | 114 | Many |
| del8p | 2.4 (12) | 21.84 | 23.19 | 1.35 | 27 | Includes *EGR3* and *DOK2* |
| gain8q | 2.4 (12) | 127.06 | 127.54 | 0.48 | 1 | *POU5F1B* |
| del14q | 2.0 (10) | 90.88 | 93.70 | 2.82 | 25 | Includes *TC2N* and *UBR7* |

(n) Number of cases with biallelic deletion of the MDR.

Supplementary Table 6. Median Survival times

| **Cohort** | **Complexity group** | **OS (years)** | **p** | **PFS (years)** | **p** |
| --- | --- | --- | --- | --- | --- |
| **CLL4** | HGC | 4.21 |  | 1 |  |
|  | LGC | 6.4 | <0.05 | 2.54 | <0.05 |
| **ARC/ADM** | HGC | 4.84 |  | 2.67 |  |
|  | LGC | 6.82 | <0.05 | 5.09 | <0.05 |

Supplementary Table 7- Univariate Cox Regression analyses in CLL4, for progression free survival (PFS) and overall survival (OS).

|  | **PFS** | | |  | **OS** | | |
| --- | --- | --- | --- | --- | --- | --- | --- |
| **Variable** | **Hazard Ratio** | **p-value** | **95% CI** |  | **Hazard Ratio** | **p-value** | **95% CI** |
| LGC | 0.72 | 0.03 | 0.54 - 0.96 |  | 0.76 | 0.07 | 0.57 - 1.02 |
| IGC | 1.20 | 0.32 | 0.84 - 1.7 |  | 1.05 | 0.8 | 0.73 - 1.52 |
| HGC | 1.49 | 0.04 | 1.02 - 2.2 |  | 1.59 | 0.018 | 1.08 - 2.34 |
| Age | 1.01 | 0.12 | 1 - 1.03 |  | 1.05 | 6.00E-10 | 1.03 - 1.07 |
| U-CLL | 2.39 | 0.00 | 1.74 - 3.27 |  | 2.31 | 2.10E-07 | 1.67 - 3.19 |
| del11q | 1.92 | 0.00 | 1.36 - 2.72 |  | 1.44 | 0.043 | 1.01 - 2.05 |
| ATM mutation | 0.96 | 0.88 | 0.53 - 1.71 |  | 1.37 | 0.28 | 0.77 - 2.47 |
| ATM biallelic loss | 1.93 | 0.14 | 0.79 - 4.7 |  | 1.73 | 0.22 | 0.71 - 4.22 |
| BIRC3 deletion | 1.92 | 0.00 | 1.33 - 2.76 |  | 1.33 | 0.13 | 0.92 - 1.94 |
| BIRC3 mutation | 1.04 | 0.92 | 0.49 - 2.21 |  | 0.89 | 0.78 | 0.4 - 2.01 |
| BIRC3 biallelic loss | 1.98 | 0.13 | 0.81 - 4.81 |  | 2.73 | 0.022 | 1.12 - 6.68 |
| Tri12 | 1.46 | 0.08 | 0.96 - 2.22 |  | 1.57 | 0.035 | 1.03 - 2.41 |
| del13q | 1.32 | 0.20 | 0.86 - 2.02 |  | 0.96 | 0.75 | 0.73 - 1.26 |
| biallelic del13q | 1.14 | 0.31 | 0.88 - 1.49 |  | 0.86 | 0.52 | 0.54 - 1.37 |
| SF3B1 mutation | 1.31 | 0.07 | 0.97 - 1.77 |  | 1.54 | 0.0061 | 1.13 - 2.11 |
| NOTCH1 mutation | 1.36 | 0.08 | 0.96 - 1.93 |  | 1.52 | 0.023 | 1.06 - 2.18 |
| TP53 dysfunction | 3.72 | 0.00 | 2.41 - 5.74 |  | 3.71 | 5.10E-10 | 2.38 - 5.78 |
| TMB (1 mutation) | 1.35 | 0.03 | 1.03 - 1.78 |  | 1.40 | 0.019 | 1.06 - 1.85 |
| TMB (2 mutations) | 1.43 | 0.06 | 0.99 - 2.07 |  | 1.85 | 0.0012 | 1.27 - 2.7 |
| TMB (≥3 mutations) | 1.73 | 0.05 | 1 - 2.98 |  | 1.72 | 0.057 | 0.98 - 3.01 |
| TL-L | 0.42 | 0.00 | 0.3 - 0.61 |  | 0.44 | 6.60E-06 | 0.3 - 0.63 |
| TL-I | 1.02 | 0.89 | 0.76 - 1.38 |  | 1.01 | 0.97 | 0.73 - 1.38 |
| TL-S | 1.81 | 0.00 | 1.38 - 2.39 |  | 1.79 | 3.70E-05 | 1.35 - 2.37 |
| i-CLL | 0.79 | 0.12 | 0.6 - 1.06 |  | 1.11 | 0.5 | 0.82 - 1.49 |
| n-CLL | 1.80 | 0.00 | 1.35 - 2.39 |  | 1.42 | 0.016 | 1.07 - 1.9 |
| m-CLL | 0.44 | 0.00 | 0.27 - 0.73 |  | 0.36 | 9.50E-05 | 0.21 - 0.62 |

**Supplementary Table 8**. Univariate Cox Regression analyses in ARC/ADM, for progression free survival (PFS) and overall survival (OS).

|  | **PFS** | | |  | **OS** | | |
| --- | --- | --- | --- | --- | --- | --- | --- |
| **Variable** | **Hazard Ratio** | **p-value** | **95% CI** |  | **Hazard Ratio** | **p-value** | **95% CI** |
| LGC | 0.69 | 0.027 | 0.49 - 0.96 |  | 0.83 | 0.48 | 0.51 - 1.37 |
| IGC | 1.10 | 0.63 | 0.75 - 1.6 |  | 0.76 | 0.38 | 0.42 - 1.4 |
| HGC | 1.98 | 0.0027 | 1.26 - 3.12 |  | 2.14 | 0.015 | 1.14 - 4 |
| Age | 1.01 | 0.56 | 0.99 - 1.03 |  | 1.04 | 0.023 | 1 - 1.07 |
| U-CLL | 2.72 | 2.60E-07 | 1.83 - 4.04 |  | 1.60 | 0.085 | 0.93 - 2.76 |
| del11q | 2.07 | 0.00068 | 1.35 - 3.18 |  | 0.67 | 0.35 | 0.29 - 1.56 |
| ATM mutation | 0.98 | 0.94 | 0.56 - 1.7 |  | 0.85 | 0.7 | 0.36 - 1.96 |
| ATM biallelic loss | 1.83 | 0.051 | 0.99 - 3.39 |  | 1.61 | 0.3 | 0.64 - 4.02 |
| BIRC3 deletion | 2.21 | 0.00033 | 1.42 - 3.44 |  | 0.98 | 0.96 | 0.45 - 2.15 |
| BIRC3 mutation | 0.53 | 0.27 | 0.17 - 1.67 |  | 1.59 | 0.43 | 0.5 - 5.06 |
| BIRC3 biallelic loss | 2.31 | 0.018 | 1.13 - 4.73 |  | 0.78 | 0.74 | 0.19 - 3.28 |
| Tri12 | 1.04 | 0.85 | 0.66 - 1.64 |  | 1.56 | 0.15 | 0.85 - 2.86 |
| del13q | 0.93 | 0.66 | 0.67 - 1.29 |  | 0.75 | 0.24 | 0.46 - 1.21 |
| biallelic del13q | 0.60 | 0.083 | 0.33 - 1.08 |  | 0.34 | 0.059 | 0.11 - 1.1 |
| SF3B1 mutation | 1.32 | 0.16 | 0.9 - 1.94 |  | 0.82 | 0.54 | 0.44 - 1.54 |
| NOTCH1 mutation | 0.92 | 0.73 | 0.57 - 1.47 |  | 1.47 | 0.23 | 0.78 - 2.74 |
| TP53 dysfunction | 4.63 | 8.20E-13 | 2.92 - 7.34 |  | 2.94 | 0.00041 | 1.57 - 5.49 |
| TMB (1 mutation) | 1.21 | 0.27 | 0.87 - 1.68 |  | 1.40 | 0.17 | 0.86 - 2.26 |
| TMB (2 mutations) | 1.48 | 0.069 | 0.97 - 2.26 |  | 0.95 | 0.87 | 0.48 - 1.85 |
| TMB (≥3 mutations) | 2.30 | 0.0035 | 1.29 - 4.09 |  | 2.47 | 0.02 | 1.12 - 5.44 |
| TL-L | 0.39 | 1.50E-08 | 0.28 - 0.55 |  | 0.42 | 0.00056 | 0.26 - 0.7 |
| TL-I | 1.80 | 8.00E-04 | 1.27 - 2.56 |  | 1.50 | 0.12 | 0.9 - 2.5 |
| TL-S | 1.92 | 0.00032 | 1.34 - 2.76 |  | 1.99 | 0.0085 | 1.18 - 3.34 |
| i-CLL | 0.85 | 0.42 | 0.57 - 1.26 |  | 0.82 | 0.51 | 0.45 - 1.48 |
| n-CLL | 2.57 | 8.00E-08 | 1.8 - 3.67 |  | 2.41 | 0.00091 | 1.41 - 4.13 |
| m-CLL | 0.31 | 1.90E-06 | 0.18 - 0.51 |  | 0.30 | 0.0027 | 0.13 - 0.69 |
