## Supplementary Methods for "High-risk Molecular Features Eclipse Genomic Complexity in Predicting CLL Patient Outcomes; Insights from the UK CLL4, ARCTIC and ADMIRE Trials"

Targeted Sequencing

Forty-one additional CLL patients were resequenced using an Agilent SureSelect XT HS2 Targeted Enrichment system (**Table S3**). Prepared libraries were sequenced on the Illumina NovaSeq 6000. QC of raw *.fastq* files was performed using FastQC, and the Mosdepth tool (v0.3.4) was used to assess sequencing coverage. After adapter trimming using Agilent’s AGeNT software (v3.0.6), FASTQ files were aligned to the hg38 (release 13) reference genome using BWA-MEMv.0.7.17. SAMTOOLS v1.2.3 converted SAM files into BAMs and Picard v2.8.3 sorted and indexed the BAMs. The CReaK module from the Agilent AGeNT software (v3.0.6) was used to mark duplicate reads and the BamUtil clipOverlap module was used to clip overlapping read pairs in the .bam files. Variant calling was performed used GATK (v4.1.9.0) and the Mutect2 (v4.1.4.1) software. After annotation, variants were filtered to enrich for high confidence somatic mutations. Initial filtering excluded intronic and intergenic variants, synonymous mutations, variants with a frequency > 1% in databases of known germline variation, and variants with a total depth less than 30. Variants were only included if previously observed as somatically acquired in CLL or annotated in COSMIC (v84). *TP53* variants were only included if they were present in the International Agency for Research on Cancer (IARC) TP53 database (vR19). *ATM* variants were included if they were present in the Leiden open variation database (LOVD) and were observed in AT families as pathogenic. Variants passing filter were manually curated in Integrative Genomics Viewer (IGV) according to the standard operating procedure described by Barnell *et al* (1).

Copy number profiling

Conumee (2) was used to produce profiles of copy number alterations (CNA) from mean intensity signals for 32 ARC/ADM patients profiled using the Illumina Infinium Human Methylation 450 BeadChip. Sub-chromosomal gains and losses were included if they were defined by a minimum of 5 consecutive probes and had segmented means of +/- 0.1 (log_2_ ratio of probe intensity) and *p*<0.001 (3).

One hundred and forty-three CLL4 patients were copy number profiled by shallow WGS (4) using the Agilent SureSelect QXT system according to manufacturer’s recommendations. FASTQ files were aligned to the hg19 human reference genome using BWA-MEMv.0.7.17, which was later converted to hg38 via liftover using the UCSC genome browser. SAMTOOLS v1.9 converted SAM files into BAMs and Picard v2.8.3 sorted and indexed the BAMs. Analysis was performed in R (v4.1.2) using the QDNASeq (v1.26.0) package (5). The data was divided into 30kb bins, normalized and corrected for mappability and GC content, log2-transformed and CNA calls were made using CGHcall (v2.58.0) (6). For sub-chromosomal CNA outside of established regions, calls had to exceed a log_2_ ratio of +/-0.2. After passing the inclusion criteria for individual technologies, all CNA were manually curated by two independent experienced researchers.

2.	Hovestadt V ZM. conumee: Enhanced copy-number variation analysis using Illumina DNA methylation arrays. R package version 1.9.0, <http://bioconductor.org/packages/conumee/>. [

3.	Marzouka	Nour-al-dain , Nordlund	Jessica	, Bäcklin	Christofer L. , Lönnerholm	Gudmar	, Syvänen	Ann-Christine	, Carlsson AJ. CopyNumber450kCancer: baseline correction for accurate copy number calling from the 450k methylation array. Bioinformatics. 2016;32(7).

4.	Parker H, Carr L, Syeda S, Bryant D, Strefford JC. Characterization of Somatically-Acquired Copy Number Alterations in Chronic Lymphocytic Leukaemia Using Shallow Whole Genome Sequencing. Methods Mol Biol. 2019;1881:327-53.

5.	Scheinin I, Sie D, Bengtsson H, van de Wiel MA, Olshen AB, van Thuijl HF, et al. DNA copy number analysis of fresh and formalin-fixed specimens by shallow whole-genome sequencing with identification and exclusion of problematic regions in the genome assembly. Genome Research. 2014;24:2022-32.

6.	van de Wie MA, Kim KI, Vosse SJ, van Wieringen WN, Wilting SM, B Y. CGHcall: calling aberrations for array CGH tumor profiles. Bioinformatics. 2007;23(7).
